# Effects of the Covid-19 pandemic on the continuity of sexual and reproductive health care in the maternity ward of a health and social promotion centre in Burkina Faso: a qualitative study

**DOI:** 10.1101/2024.04.10.24305634

**Authors:** Patrice Ngangue, Mariam Ahmat Mahamat, Danièle Sandra Yopa, Gbètogo Maxime Kiki, Nestor Bationo, Douglas Mbang Massom, Josiane Seu, Birama Apho Ly

**Author notes:** **Correspondence:** Patrice Ngangue, MD, MSc, PhD, Adjunct Professor Faculty of Nursing Sciences, Laval University, 1050 avenue de la médecine, Quebec, QC, G1V 0A6, Canada.

## Abstract

**Background:** The COVID-19 pandemic has significantly impacted the continuity of maternity care in Burkina Faso. This study aimed to compare the volumes of in-person visits and to explore the experiences of healthcare providers and users regarding the continuity of healthcare in the maternity ward of a Health and Social Promotion Center (HSPC) before, during and after the lockdown during the COVID-19 pandemic in Ouagadougou, Burkina Faso.

**Methods:** We conducted a multimethod, cross-sectional exploratory study with a phenomenological approach. Monthly health administrative data regarding family planning visits, antenatal visits, deliveries, and postnatal care before, during, and after COVID-19 were collected and compared. Qualitative data were collected through semi-structured interviews with family healthcare providers and users and thematically analyzed.

**Results:** The study found that the pandemic led to a decline in the demand for healthcare, as people were afraid of contracting COVID-19. This was particularly true for pregnant women who had recently given birth. The study also found that the pandemic disrupted the supply of essential medicines and medical supplies, which made it difficult for healthcare providers to provide quality care.

The qualitative analysis allowed us to highlight three themes: the representation of respondents on COVID-19, their perception of the effectiveness of barrier measures and their analysis of the continuity of care in times of COVID-19: the picture of respondents on COVID-19, their perception of the efficacy of barrier measures and their analysis of the continuity of care in times of COVID-19. Despite these challenges, the study found that healthcare providers and users could find ways to maintain continuity of care.

**Conclusion:** The study concludes that the COVID-19 pandemic has significantly impacted the continuity of maternity care in Burkina Faso. However, healthcare providers and users have found ways to maintain continuity of care, and the study provides recommendations for improving continuity of care in the future.

## Introduction

In 2020, the COVID-19 pandemic profoundly impacted the world beyond expectation. Statistics reveal that at least 135 million people were infected, with more than 2.9 million deaths. As a result, the scourge has hurt the normal functioning of many health systems. Also, the lockdown has led to a deterioration in access to health care for the less privileged and most vulnerable populations. The lockdown has led to a decline in access to health care for the less privileged and most vulnerable people [1, 2]. Since the declaration of the first case of coronavirus disease, a drop in the use of health services has been observed in most health facilities, which risks destroying the efforts made so far and increasing the number of extra-hospital deaths of groups related to other diseases [3]. Indeed, non-emergency health care has been relegated to the background to prioritize the care of thousands of patients with COVID-19 [4]. However, some services, such as maternal healthcare and family planning, cannot be put on hold [5].

Continuity of sexual and reproductive health care is one of the key elements of quality service. It designates how patients perceive their services as a succession of related events, coherent and compatible with their needs and personal situations [6]. Based on the recommendations of the World Health Organization, measures must be planned to safeguard essential preventive health care and care for the most vulnerable groups, including women, children and youths [7].

Like other countries worldwide, Burkina Faso has been heavily affected by COVID-19. The scourge has set in motion an already fragile and poorly organized health system, with repercussions on women and girls’ access to reproductive health services, particularly family planning [1, 8]. There was a disruption in the supply chains of contraceptive products, the closure of health planning services and increased women’s fear of going to health facilities [9, 10].

Given this situation, the continuity of essential health care within health facilities, particularly those related to mothers, children and adolescents during the management of a pandemic, was highlighted [11, 12]. However, little data on the impact of the lockdown on sexual and reproductive health are available to date. Therefore, this study analyses patients’ perceptions of said services and clinical data during different periods around the lockdown. The objective was to understand the effects of the COVID-19 pandemic on the continuity of maternity care at the Wemtenga Health and Social Promotion Center.

## Material and methods

### Study design

We conducted a multimethod, cross-sectional exploratory study with a phenomenological approach. The phenomenological approach involves observing and describing phenomena, and their modes of observing and describing phenomena and their methods of appearance are considered independently of any value judgment [13]. Multimethod research may be broadly defined as employing two or more different methods or styles of research within the same study or research program rather than confining the research to a single method [14]. In this study, the documentary review was used to explore and analyze the experiences of healthcare providers and users on the continuity of healthcare in the maternity ward of the Health and Social Promotion Center (CSPS) in Wemtenga at different times of the COVID-19 pandemic. that is, before, during and after the pandemic.

### Participants and recruitment

The study was conducted on reproductive healthcare providers (midwives and birth attendants) and clients benefiting from this reproductive healthcare of the maternal and child care and family planning and maternity services of the CSPS of Wemtenga. Systematic sampling was conducted to recruit healthcare providers, while client recruitment was done by reasoning sampling. Recruitment was done gradually until the saturation of information.

To be included, healthcare providers should have been state-qualified midwives and mæutician and have experience of at least three years in the service at the time of the study. As for clients, they should have been beneficiaries of Reproductive Healthcare (RH) and have made at least one visit to one of the RH services during or after the lockdown. Excluded from the study were all providers and all beneficiary clients known to the research team.

### Data collection

To collect the data, we used semi-structured interviews and documentary reviews. The choice of the semi-structured interview is justified by its flexibility for the participant and the ease it offers the researcher to explore all the themes related to the research problem. The interview guide consisted of questions structured around Bachrach’s dimensions of continuity of care to explore the different research questions [15].

The documentary review consisted of objective information to understand better the effects of the COVID-19 pandemic on the continuity of care at the CSPS Wemtenga maternity hospital. All documents related to the continuity of care during this pandemic (consultation registers, hospitalization registers, health books or pictures associated with the continuity of healthcare during COVID-19) were used. The information collected was triangulated with the interviews. It was carried out using a grid containing questions about the number and the reasons for visits to understand the effects of the COVID-19 pandemic on the continuity of care at the Wemtenga CSPS maternity ward.

### Data analysis

Individual interviews were recorded, listened to several times and then fully transcribed. The verbatim was coded using Nvivo 12 software. A thematic analysis was carried out using the Braun & Clarke Model (2006). The exhaustive codification of all the verbatims was carried out by classifying them by addressed themes (the representation of COVID-19, the effectiveness of barrier measures and the analysis of continuity of healthcare). After categorizing codes into sub-themes and then into general themes, they were organized by a unit of meaning, making it possible to understand the effects of the pandemic on the continuity of healthcare. Finally, the themes were reviewed and refined. Lincoln and Guba’s qualitative research scientific criteria were ensured throughout the study. These criteria were credibility, transferability of the conclusions, confirmability and reliability.

The analysis of healthcare demand before, during, and after the COVID-19 pandemic aims to enhance understanding of the pandemic’s effects on the continuity of healthcare services at the maternity ward of the WEMTENGA CSPS compared to the verbatim statements recorded. Data regarding healthcare demand before, during, and after the lockdown were collected from the records of different services and then encoded in Microsoft Excel 2016. Subsequently, these data were analyzed using the Epi Info 7 software. The results were presented in tables to better visualize variations in healthcare demand before, during, and after the lockdown. The percentage points (pp) were calculated to describe the variations between the percentages of healthcare demands between the periods.

### Ethical considerations

The study obtained authorization from Burkina Faso’s Ethics Committee for Health Research (CERS) under number 2021-12-284. In addition, administrative approval was delivered by the regional health directorate. All study participants gave informed consent to the study by signing an information and consent form before participating in the surveys. They were free to withdraw from the study at any time without prejudice. Data confidentiality was ensured by assigning a code to each participant. The interviews were individual. The place and date of the interviews were at the participant’s convenience.

## Results

### Sociodemographic characteristics of participants

A total of 30 participants were enrolled in our study (5 healthcare providers and 25 patients). The health care providers were women aged 35 to 44 years. The age groups represented the most were those aged 25 to 34 and those aged 35 to 44, hence 36% each. All healthcare providers had a higher level of education, while patients were primarily uneducated (32%) (see Table 1). Of the five (5) healthcare providers interviewed, three (3) lived more than 5 km from the healthcare centre. On the other hand, all the patients lived less than 5 km from the healthcare centre.

**Table 1:** Sociodemographic profile of participants.

| Terms |  | Care providers |  | Patients |  |
| --- | --- | --- | --- | --- | --- |
|  |  | Number (n) | Percentage (%) | Number (n) | Percentage (%) |
| age range | 15-24 years old | 00 | 00 | 07 | 28 |
|  | 25-34 years old | 00 | 00 | 09 | 36 |
|  | 35-44 years old | 05 | 100 | 09 | 36 |
|  | ≥45 years old | 00 | 00 | 00 | 00 |
| Study level | Uneducated | 00 | 00 | 08 | 32 |
|  | Primary | 00 | 00 | 03 | 15 |
|  | Secondary | 00 | 00 | 07 | 28 |
|  | Superior | 05 | 100 | 07 | 28 |

### Presentation of documents used for the documentary review

For our study, we used several sources of information, including consultation registers and patient files.

### Results of interview analyses

The data analysis allowed us to highlight three themes, namely: the representation of respondents on COVID-19, their perception of the effectiveness of barrier measures and their analysis of the continuity of care in times of COVID-19: the picture of respondents on COVID-19, their perception of the efficacy of barrier measures and their analysis of the continuity of care in times of COVID-19.

### Picture of COVID-19

For most participants, COVID-19 is a contagious and fatal disease, as indicated by the statements of some providers:

“Covid-19 is a contagious respiratory disease which is fatal, especially for whites (…). It seems that it can’t stand our sun until then. That’s why we die less than white people. Otherwise, it kills a lot” [Ps2].

“Ah, COVID there is a contagious disease that kills people (…) we saw on TV how many people died from that among white people” [P19].

Virtually all the participants associated COVID-19 with cough, fever, chills and headache, reinforcing their fear of having the disease when they were in front of those with one of these symptoms. This led to the acceptance of the use of barrier measures, as indicated in the following quote:

“We learned that it manifests itself in coughs, headaches, hot body and when you have trouble breathing (…); that’s why when we went somewhere, and someone started coughing strangely or when he sweats a little too much, we were afraid that he would give us the disease” [P8].

Almost all participants cited social distancing, washing hands, wearing a mask and coughing into the bend of the elbow as barrier measures. Moreover, most believe these measures are more effective if everyone applies them. However, many lamented the discomfort generated by the permanent wearing of the face mask, which makes breathing suffocating in most cases. In addition, for most healthcare providers, it was painful to apply social distancing when providing patient care.

“Yes, those barrier measures are good because they help us not to get infected with the disease, but face masks. It’s not easy. I can’t breathe in it (smile)” [P3].

“I find the barrier measures effective because we cannot transmit the disease to each other (…), but when we have to do the consultation, it is difficult because we cannot communicate well with the patients…” [Ps1].

Also, healthcare providers mentioned the lack of personal protective equipment, which sometimes made them feel insecure about the disease, as described below:

“When COVID arrived, we saw dead people so much that we were afraid, so we did everything to respect the barrier measures there, but it must be said that we were not given face masks all the time here. At the start of Covid, we had to buy them ourselves sometimes because we were afraid, but afterwards, it was better; we had the masks and gels, and there was a corner for people to wash their hands " [PS2].

### Analysis of Continuity of Care in Times of COVID-19

#### Availability of care

The CSPS ensured the continuity of maternal and child health care despite the lockdown. In addition, the care services offered are of better quality thanks to the systematic application of barrier measures by both patients and caregivers.

“For care, nothing has changed; it’s still the same thing. When you arrive at the hospital, you feel that everything is clean, and that’s good. We feel safe in any case” [P23].

Yet patients described feeling underinformed about COVID-19. They lamented the lack of communication on the disease within the health structures. Their source of information was their entourage and the media.

“During COVID, we heard things on the radio and TV but did not know what was true or false. Even when you come to the hospital, you don’t dare to ask, and the doctors didn’t tell us anything” [P19].

In fear of contracting the disease and concerned about protecting their loved ones, the professionals interviewed declared that they had been more vigilant about compliance with barrier measures during care services during the pandemic than before the pandemic.

“We were afraid of contaminating ourselves and bringing it home; we were cautious. Sometimes, some women forget to wear face masks, and we have to remind them of that because we treat people well, but we must also protect ourselves” [Ps4].

#### Accessibility to healthcare

According to the participants, the accessibility to healthcare has been altered not because of the distance between the centre and the patients’ homes but rather because of the reduction in the reception capacity of the maternity ward due to the social distancing work.

“… as we have to put 1 meter between each patient, we didn’t have enough space for everyone. If we had to receive the same number of patients as before, we wouldn’t be able to” [Ps1].

“…there are times, even if you have an appointment, you have to come early; otherwise, when you come a little late, there is no more room because of that 1-meter distance, and you have to go back home” [P8].

Also, patients and companions without face masks were turned away, as evidenced by the words of a patient.

“Once, I came to take my pill and forgot to wear the face mask. The security guard let me in, but the midwife told me to go out and get it when I entered the room. If not, she can’t take me” [P15].

### Request for healthcare

The respondents affirm that the drop in the demand for healthcare between the periods before and after the lockdown could be explained by the fear of contracting the disease or the constraint of having to wear a face mask during the hospital stay, as this patient explained:

“If it’s not because of that illness; I didn’t go to the hospital huh (…) when you come you have to wear that face mask all the time (…) you can’t also take it off because as it’s in the hospital, you can catch covid there because you don’t know what the other patients who are there with you are like” [P23].

Care providers, for their part, admit that this reduction in the demand for care has made it possible to reduce their workload, which has made it easier to manage the stress linked to the fear of contracting the disease and contaminating their loved ones.

“As the patients fear the disease, they no longer came too much like that (…). It helped us a lot because we had less work and could afford to provide care by being more careful” [PS4].

### Evolution of the demand for healthcare

The table below summarizes the evolution of the demand for healthcare during the first trimester before the lockdown, during the lockdown and the first trimester after the end of the lockdown. It results from the documentary review that was carried out within the CSPS.

Overall, we observe a decline in healthcare demand both before and during the lockdown, followed by an increase in demand during and after the lockdown. Specifically, we noted a decrease of 6.35 and 5.56 percentage points (pp) in demand for family planning and prenatal care, respectively. The total number of healthcare requests decreased from 5,292 before the lockdown to 3,528 during the lockdown. Subsequently, after the lockdown, the number of requests increased to 4,787. The percentage points (pp) increased to 6.02 and 4.65, respectively, for family planning and prenatal care after the lockdown, while during the lockdown, they stood at [specific value needed] for both. Antenatal care consistently remained the primary demand within the maternity ward throughout the various periods examined in the study. We noticed maternal and child healthcare demand decreased sharply before and during the lockdown. For example, the need for family planning care fell almost by half, dropping from 1596 to 840 between the two periods. However, the exception is made with deliveries where there is an increase in demand. We go from a monthly average of 280 to 308 deliveries between the two periods.

After the lockdown, there has been a rise in the demand for healthcare in general, which remains, however, lower than the demand before the lockdown. Conversely, there is instead a regression in childbirth during the post-lockdown period. We go from 924 deliveries to 839 after the lockdown (see Table 2).

**Table 2:** **Sociodemographic** profile of participants: demand for healthcare before, during and after the lockdown.

| Terms | T1= Request for care during the first trimester before the lockdown |  | T2= Request for care during the lockdown |  | T3= Request for care after the first trimester after the end of the lockdown |  | PP= T2-T1 | PP= T3-T2 |
| --- | --- | --- | --- | --- | --- | --- | --- | --- |
|  | N | % | N | % | N | % |  |  |
| Planning family care | 1596 | 30.16 | 840 | 23.81 | 1,428 | 29.83 | -6.35 | 6.02 |
| Prenatal care | 2,184 | 41.27 | 1,260 | 35.71 | 1,932 | 40.36 | -5.56 | 4.65 |
| deliveries | 840 | 15.87 | 924 | 26.19 | 839 | 17.53 | 10.32 | -8.66 |
| Postnatal care | 672 | 12.70 | 504 | 14.29 | 588 | 12.28 | 1.59 | -2.01 |
| Total | 5,292 | 100 | 3,528 | 100 | 4,787 | 100 |  |  |

## Discussion

With the advent of COVID-19, essential health services such as reproductive and family planning have been disrupted in most parts of the country, obliging health authorities to mobilize all available resources to cope with this pandemic. Our study aimed at understanding the effects of COVID-19 on the continuity of maternity care at the WEMTENGA Health and Social Promotion Center (CSPS).

Our results revealed that the availability of maternity care was effective, with improved quality of care offered. To limit the spread of the disease, the staff had become more alert and respected the barrier measures during their services. Accessibility to care was reduced during the lockdown due to reduced reception capacity within the maternity ward and social distancing.

The documentary review enabled us to observe a decrease in the demand for care before and during the lockdown and a resurfacing of the need for care during and after the lockdown. Between the period before and during the lockdown, the demand for maternal and child health care fell sharply. This is supported by a study by Dell’Utri et al., which compared obstetric and gynecological emergency care admissions during COVID-19 with those of the previous year and found a decrease in admissions of approximately 35% [16]. Also, a systematic review showed that all countries experienced reduced provision and use of services [2]. Another study of sexual healthcare professionals in the USA indicates the service management of abortions and testing for HIV and sexually transmitted infections has been reduced by 76%, 75% and 82%, respectively [17].

### Accessibility to healthcare

Our study revealed that health negatively affected care accessibility at the CSPS. While social distancing motivated the limitation of the number of nursing staff and patients interfering together, it simultaneously worked to reduce the accessibility of care in the service. The reception capacity was reduced, and wearing a face mask was also a major constraint for patients who wanted to access the service. The documentary review confirmed the extent of the phenomenon and revealed a significant drop in the care requested during the lockdown compared to that formerly requested before the lockdown. The data from our study reflects a reality faced by many health systems worldwide, such as in China [18]. A study conducted in Uganda reveals that during the four weeks of the lockdown, all antenatal services, vaccination and sexual health services were closed entirely from March 23 to April 21, 2020 [19].

### Request for healthcare

The decrease in the demand for healthcare between the period before and after the lockdown can be attributed to the difficulties linked to the accessibility of care, but also to the fear of contracting the disease, infecting relatives and dying is explained by the fear of contracting the disease and the constraint posed using the face mask. This drop-in activity for healthcare providers has benefited them, thus limiting work exhaustion, stress and the risk of contamination. An ecological study using a national insurance database in France showed that prescriptions for contraceptives and ovulation indicators initially increased by 47% and 16% during the first two weeks of the lockdown, but that they then decreased considerably. This decrease is [20]. The fear of disease is also one of the main reasons for giving up, as mentioned by the users surveyed in France. This survey shows that 39% of respondents expressed anxiety about COVID-19 [21].

Health professionals have seen a significant decrease in the use of care unrelated to COVID-19 in the city and establishments during the first lockdown. People suffering from illnesses have given up treatment for fear of being contaminated or considering their health condition secondary to COVID-19 patients [22]. In a systematic review, the included 81 studies reported 143 estimates of changes in healthcare utilisation between pandemic and prepandemic periods, of which 136 (95.1%) were a reduction [23]. In the mental health field, a study conducted in Korea reported that medical service use decreased overall, with a particularly significant decrease in emergency departments. As the pandemic worsened, the decline in outpatient visits became more pronounced among those with severe mental illness [24].

Nevertheless, even if the data tend to show that this impact was the same throughout the world, several countries have been able to implement alternatives to the administration of direct care, which has made it possible, despite the fears and problems linked to accessibility, to continue to monitor patients. In France, to deal with the isolation or the impossibility of certain people accessing healthcare services, teleconsultation was facilitated during the lockdown and covered 100% by insurance. Telehealth has experienced rapid growth, thus facilitating care and reassuring patients and their families [25]. Only deliveries have not been altered. This can be explained by the fact that not only can deliveries not generally be scheduled or delayed, but care benefits from free care and is much less risky in a hospital setting than in a community setting. As a result, defying the fear of illness to protect the life of the future mother and her child is much easier.

For decades, most countries have not faced a tragedy like the COVID-19 pandemic. Through this work, we identified, in the case of the CSPS in Burkina, the health system’s flaws and difficulties in ensuring the maintenance of essential services. We believe it is more than urgent for health systems to show more resilience. Therefore, this study should be strengthened, and this type of study should be extended to other services and the whole country to assess the gap caused by the pandemic.

### Strengths and limitations of the study

The main strength of this work is the in-depth exploration of the factors that influence the continuity of healthcare during a pandemic through the combination of the qualitative approach with the documentary review.

This study is not without limits. The subjectivity of the researchers probably impacted the results during the data analysis. However, the codes were approved by the research team. Additionally, the study stopped to assess the effects of COVID-19 at a single centre level. The results are only transferable in the context of Wemtenga’s CSPS.

## Conclusion

COVID-19 is a global pandemic that has plunged all countries into a crisis whose effects have been felt not only in health, economic and social terms. At the health level, the growing increase in cases and the high mortality rate surprised the health systems, which were not sufficiently prepared to face such a threat. Aware of this weakness, the research team proposed to conduct the present study on the effects of COVID-19 on the continuity of maternity care at the Wemtenga Health and Social Promotion Center (CSPS). It revealed an improvement in the quality of care provided since the advent of Covid-19 thanks to a more docile attitude of patient users. However, there is also a decrease in the use of this service during the lockdown. This is explained by users’ fears of contamination and the constraints linked to applying barrier measures, which negatively influence care accessibility. On the strength of these different results and not claiming any exhaustiveness, our service could serve as a basis for other scientific studies. So, we’d suggest that actions be taken to ensure the best care of future health crises.

## Data Availability

All data produced in the present work are contained in the manuscript

## Author Contributions

All authors contributed equally to this work. All authors agree to be accountable for the content of the work.

## Acknowledgments

We would like to thank all the healthcare staff for their availability and support, without which this work would not have been possible. We are also grateful to all the participants and all those who have directly or indirectly contributed to the smooth running of the survey.

